# An Immune System for the City: A Randomized Trial of a New Paradigm for Surveillance and Control of Disease Vectors

**DOI:** 10.1101/2024.05.30.24308159

**Authors:** Michael Z Levy, Laura D. Tamayo, Carlos E. Condori-Pino, Claudia Arevalo-Nieto, Ricardo Castillo-Neyra, Valerie A. Paz-Soldan

## Abstract

Vector-borne pathogens continue to emerge, kill and harm humans with unrelenting regularity. Conventional strategies to controlling insect vectors grew out of the military; communication is hierarchical, responses unilateral, and regulation predetermined. We developed an alternative approach, modeled after the adaptive immune system, and compared the approaches through a cluster-randomized trial in the context of an ongoing urban Chagas disease vector control campaign in Arequipa, Peru. We report here early results from a pre-planned interim analysis. In the intervention (immune) arm 23 infested households were detected in 10 separate foci; in the control arm only 5 infested households were detected, and all were from the same focus. The immune approach was adaptive, and more effort was expended upon detection of an infestation (1085.2 person days in the immune arm vs 864.2 in the conventional; Rate ratio 23/1085.2:5/864.2 = 3.66 [1.35 12.38], p-value =0.0062). Vector surveillance approaches modeled after the immune system may be more effective than conventional approaches, especially in cities and other complex civilian environments.

**Author summary:** Vector-borne diseases kill and harm millions of people each year; many more infections are prevented through vector control campaigns. These have been developed in military contexts, and their hierarchical structures may be less effective in civilian settings. We developed an alternative approach to vector control, one that drew inspiration from the adaptive immune system, and show that it outperformed the conventional approach in the context of an ongoing Chagas disease vector control campaign in the city of Arequipa, Peru. Vector control strategies must work across scales, be flexible to shifting aims and budgets, and effectively harness the strengths of the communities they purport to protect. The immune system provides an effective model to achieve these aims.

## Introduction

The difficulties faced by a community when trying to control a dangerous insect are in many ways analogous to those faced by the body when attacked by a pathogen. Through evolution, mammals have developed a complex and interlocking defense against invaders—an immune system well-honed to detect and control infections. The adaptive immune system relies on communication among autonomous entities to recognize pathogens and recruit effector cells (1); it includes a combination of systemic and local responses to remove pathogens; and, most amazingly, tightly regulates these responses and creates memory to improve them in the future. (2)

Conventional strategies for vector control, by contrast, grew out of the military; with hierarchical communication, unilateral responses, and predetermined regulation. (3) Despite their ubiquity, top-down control campaigns have rarely been challenged, formally, by alternative approaches. We conducted a cluster-randomized trial evaluating a new paradigm for the control of dangerous insects patterned after the adaptive immune system against a conventional top-down system in an ongoing Chagas disease vector control program in the city of Arequipa, Peru. Here we report the early results from a pre-planned interim analysis.

## Materials and Methods

### Study System

Chagas disease, caused by the parasite *Trypanosoma cruzi*, is one of the principal infectious causes of morbidity and mortality in the Americas. (4,5) Since 1991, *Triatoma infestans*, the principal vector of *T. cruzi* in South America, has been the target of an elimination program known as the Southern Cone Initiative. This initiative, formalized in the early 1990s during a period of return to democracy in many countries in the region, nevertheless retained conventional top-down vector control approaches that had proven successful under more authoritarian regimes. By many measures, (6,7) the initiative has been a tremendous success, with Chile, (8) Brazil, (9) Uruguay, (10) and Paraguay (11) achieving disruption of transmission of *T. cruzi* by *T. infestans*. Peru did not initially join the Southern Cone Initiative, and control efforts in the country lagged for many years. (12) *T. infestans* is limited to Southern Peru, where it infested Arequipa, a regional capital of over one million inhabitants. (13)

Initially with support from the Canadian International Development Agency and technical support from the Pan American Health Organization, the Peruvian Ministry of Health conducted insecticide treatment of over 80,000 houses, preventing significant mortality and morbidity. (14,15) The campaign, like many conventional campaigns, (16,17) consisted of three phases: the preliminary *planning phase* which served to define the geographic areas in need of control, (18) the *attack phase* (19) during which brigades of vector control technicians applied pyrethroid insecticides to domestic and peri-domestic areas of all participating homes (twice at a target 6-month interval), and the post-spray *surveillance phase*, in which households are monitored for insects through a combination of passive and active methods.

The attack phase in Arequipa lasted from 2003 to 2018 and proceeded on a district-by-district basis. Districts entered the surveillance phase in a rolling manner, six months following completion of insecticide application. Initially, post-spray surveillance activities were mostly limited to passive surveillance, with occasional active search for infested households when personnel were available. (20,21) A series of data-driven surveillance schemes were piloted in multiple districts of the city. (14,22,23) These studies included search for the insect in over 8,000 households based on data collected during the attack phase and new data that accumulated during the surveillance phase. Only three infested households were detected through these efforts [Unpublished data].

### Intervention design

*The immune approach*: We adapted aspects of the immune system from the scale of cells to that of landscapes, incorporating these into the ongoing Chagas disease vector surveillance activities. Our analogy is described fully in Figure 1. The immune approach is the intervention arm. *The conventional approach* is an augmented version of standard practice, including traditional communication techniques and passive surveillance, along with state-of-the-art, though top-down, active surveillance methods. The conventional approach is implemented in the trial as the control arm. We outline the key ways in which the two approaches diverge, ideologically as well as methodologically, below.

**Figure 1.**
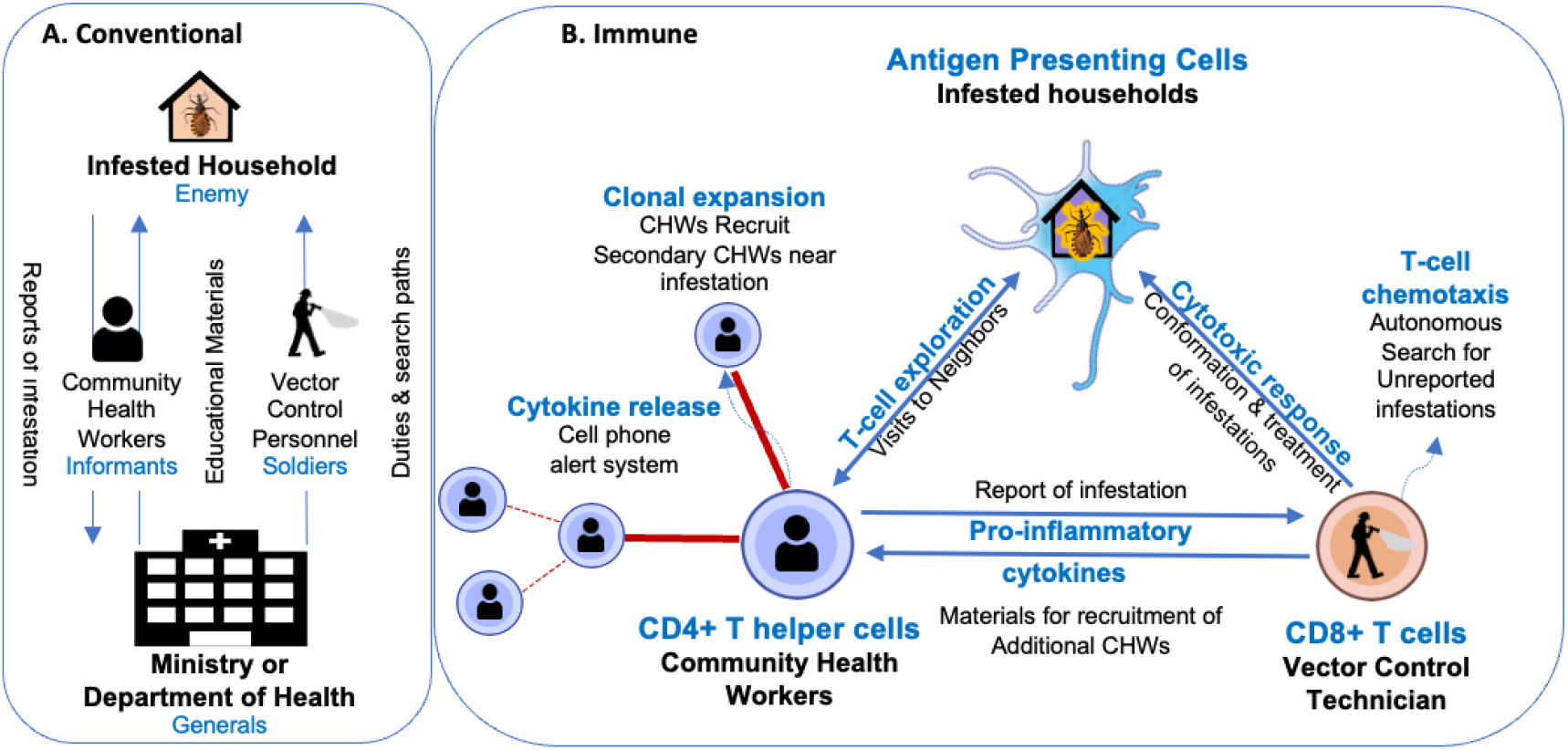
Analogies for vector control a) the conventional, military analogy, b) the immune system analogy. CHW=Community Health Worker

### Reporting

The immune system is extraordinarily egalitarian. Any of a large population of cells may present an antigen and trigger the system to act, thereby catching an invading pathogen early, before it spreads.

Under the conventional system, infested households are considered, not as antigen-presenting cells that will seek the attention of the surveillance system, but as the ‘enemy’, which will avoid it. Public health officials worry homeowners might be hiding infestations, or falsely reporting infestations in order to obtain free insecticide treatment for other nuisance insects. The lack of trust manifests in the regulations for both reporting and search (the latter is discussed below). For reporting, if a homeowner finds a triatomine they are required to capture it (24) and bring it to a primary-level health facility for identification. The requirement produces a barrier for those homeowners who are hesitant or unable to trap insects. It also presents logistical challenges as the health facility must have staff on hand to identify the insect, or to get the insect to the right person in a timely manner. Moreover, efficient data management systems are necessary to convey the identification up the chain of command to the central vector control office. Additional delays may occur in the central office where the authority and resources to program treatment of the infested house sits.

In the immune arm, we dropped the requirement to bring an insect to the health post for identification. Instead, we accepted reports by phone call, social media, text message, and Whatsapp through an integrated system we publicized as ‘AlertaChirimacha’, (25) named after the local name for the triatomine, Chirimacha. We requested, but did not require, photographs of the insects.

### Search

The immune system is decentralized; T-cells guide their own movements, aided by information imparted to them by cytokines, in much the same way that animals search their environment for food. (26) There is no control center; no physiological structure directs the T-cells. Conventional vector surveillance paradigms, by contrast, are top-down, with a central authority directing the activities of vector control personnel. We used two apps, both based on the same spatio-temporal risk models. (22) The control app assigned households for inspection to personnel, providing alternates as needed. The immune app, called VectorPoint, provided risk estimates but allowed inspectors to use their own judgment when searching their areas for insects. (14,23)

### Interaction with Homeowners

T-cells elicit information from the somatic cells they surveil, they do not enter them. Under the conventional system, vector control personnel are instructed to enter and fully inspect the premises of the households they are assigned. The requirement stems, in part, from the suspicion that householders may be hiding infestations that could be uncovered through a thorough inspection. In the immune arm we did not require vector control personnel to enter households. Instead, they were instructed to visit houses, speak to residents about Chagas disease and triatomines, and provide materials indicating how these could be reported via AlertaChirimacha. Inspectors offered to inspect households if a resident feared they might have an infestation, or if they were unable to check for insects themselves.

### Response

The control arm of our trial continued the practice of inspecting the immediate neighbors of each household found to be infested with triatomines. In the immune arm, the system was activated, temporarily, following detection of an infested household. Vector technicians were recruited from nearby areas to the affected site, which was roughly 200 meters around the infested house. Along with the original technician, they distributed flyers to all households and placed posters in local shops. In addition, Facebook posts were targeted to the area for 10 days (the radius of Facebook’s targeted posting is defined by the number of users in the area, but was typically around 1 km) and verbally by including the name(s) of the neighborhood(s) affected. The posts encouraged residents to inspect their homes for triatomines and report any suspected bug, or its traces (feces or exoskeletons), via WhatsApp, text message or phone call. If additional infested houses near the original focus were found, the radius was expanded around the new infestation, and additional inspectors recruited as necessary. A follow-up post was released on Facebook on days 10 and 30 after the initial post. Subsequent posts informed neighbors that more houses had been found positive and asked them to stay alert to the presence of triatomines and report them if found. Once all households in the affected area had been visited by technicians, those who had been recruited to the site were allowed to return to their own catchments. We posted a ‘downregulation’ message on Facebook once all infested houses were treated and 30 days had passed with no additional infestations. This message explained that the affected houses had been treated with insecticide and the insects were controlle

### Deviations from the protocol due to ‘Immune suppression’ during COVID-19

The original protocol included significant roles for community health workers (CHWs). In the conventional arm, these individuals were meant to visit their neighbors regularly to ask if they had chirimachas and, in the case of an affirmative, report the household to the corresponding health center. In the immune arm, the role of community health workers was patterned after that of CD4+ helper T-cells. They were meant to visit their neighbors, and, in the case of a neighbor reporting an infestation, communicate, via Whatsapp, directly to the vector control technician. The technician would then come to the site and activate the response, if necessary, as described above. Most importantly the community health worker was meant to ‘clonally expand’ by training others in their neighborhood to serve as community health workers, temporarily, while the immune system was activated. These secondary community health workers would then form part of the memory of the system, available for re-activation if another infestation were to appear in the area in the future. During the pandemic, the community health worker system broke down and these planned activities were dropped.

### Trial Design

We divided the area of the city under vector surveillance into 60 surveillance catchments, following existing geo-political boundaries (locality, district) wherever possible. We pair-matched these catchments (30 in the intervention and 30 in the control arm) using non-bipartite matching (R package “nbpMatching”). (27) We balanced the assignment of members of these pairs to the arms of the trial based on three criteria that we have previously found to be indicators of the risk of infestation (21): the total number of households, the number of infested houses detected in prior surveillance activities, and the years since insecticide application during the attack phase. The average sizes of catchments in the immune and control arms were 2275.7 and 2266.7 households, respectively. In the immune arm an average of 1267 houses had been treated during the attack phase in each catchment, and an average of 98.5 were found to be infested at the time of treatment. In the control arm an average of 1143.8 households were treated in each catchment, and the average number of infested households was 94.6 (Table 1). The trial is registered in socialscienceregistry.org (trial 10985).

**Table 1.**
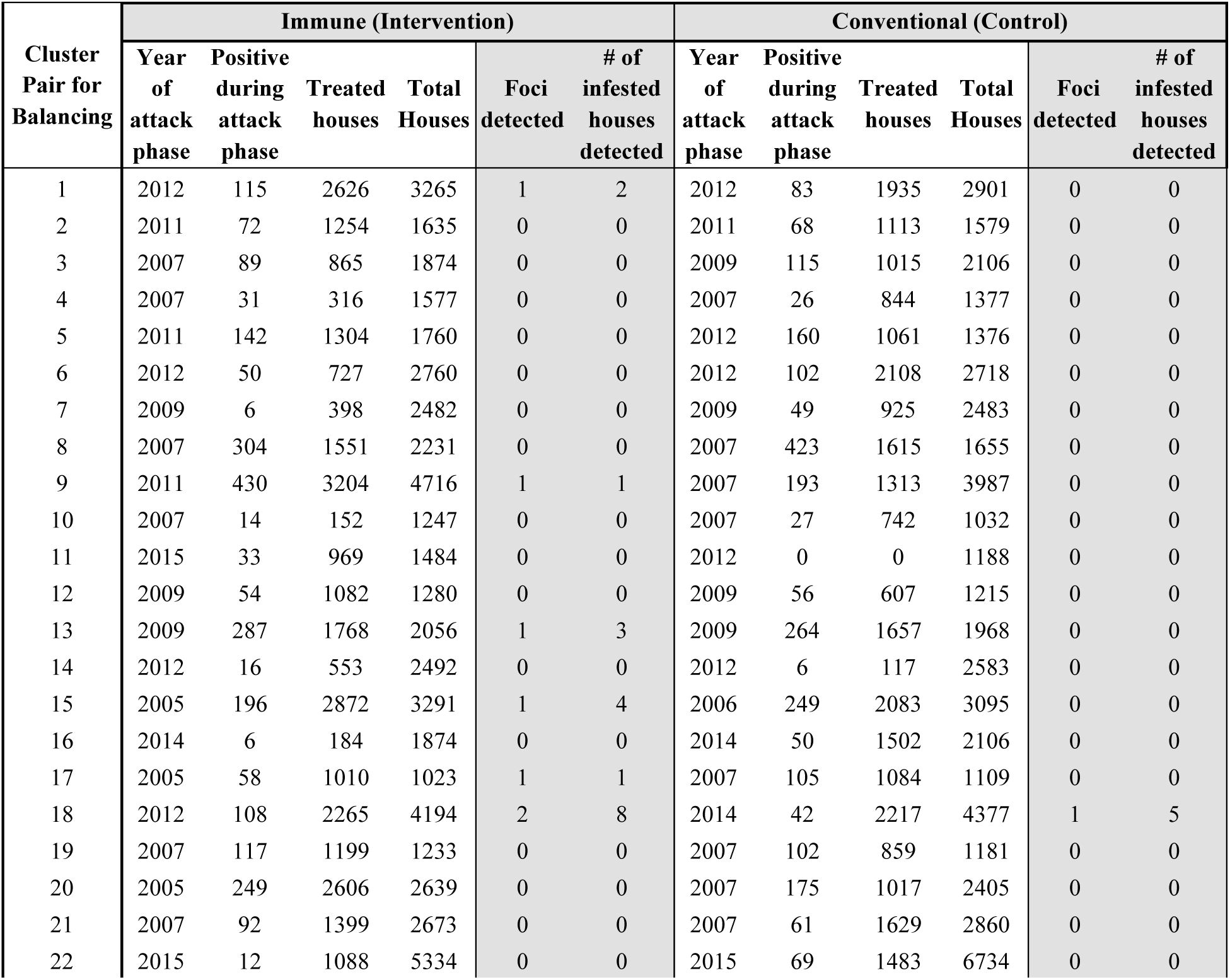
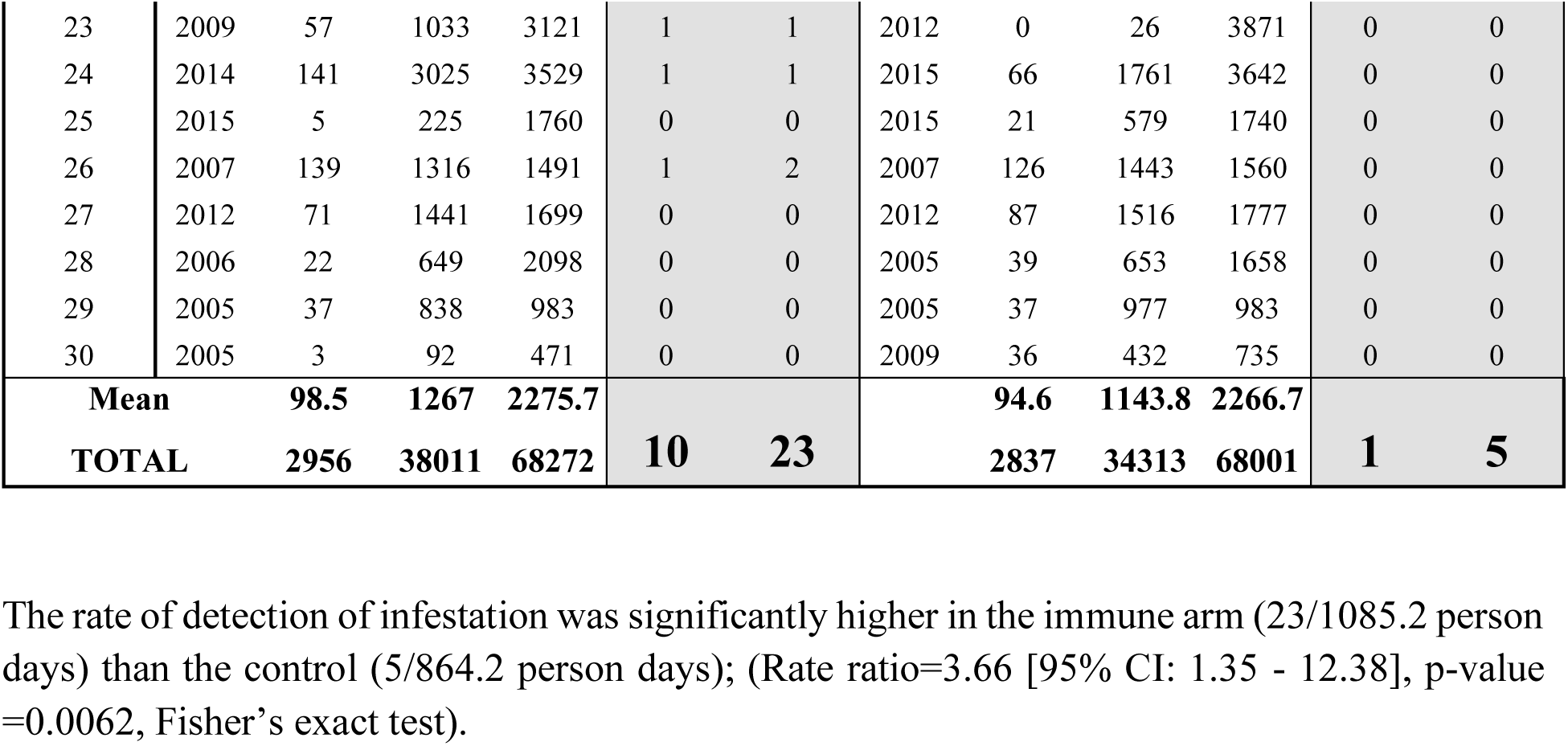
Baseline characteristics of intervention and control surveillance catchments as measured during the attack phase of the Chagas disease vector control campaign in Arequipa, Peru (white). Interim results from year 3 of a head-to-head trial of the Immune and Conventional (Control) surveillance systems (grey).

### Results

The immune approach uncovered 23 infested households, ten from the initial detection and an additional 13 during the activation of the system in the affected areas. The ten initially detected came from nine separate clusters. Five were initially reported through the AlertaChirimacha system (via Facebook or Whatsapp). Three were uncovered through active search by vector control technicians, and two were reported to health posts. The conventional approach uncovered a total of five households in a single cluster; one household brought an insect to a health post and the remaining four were immediate neighbors to the index house (Fig. 2; Table 1).

**Figure 2.**
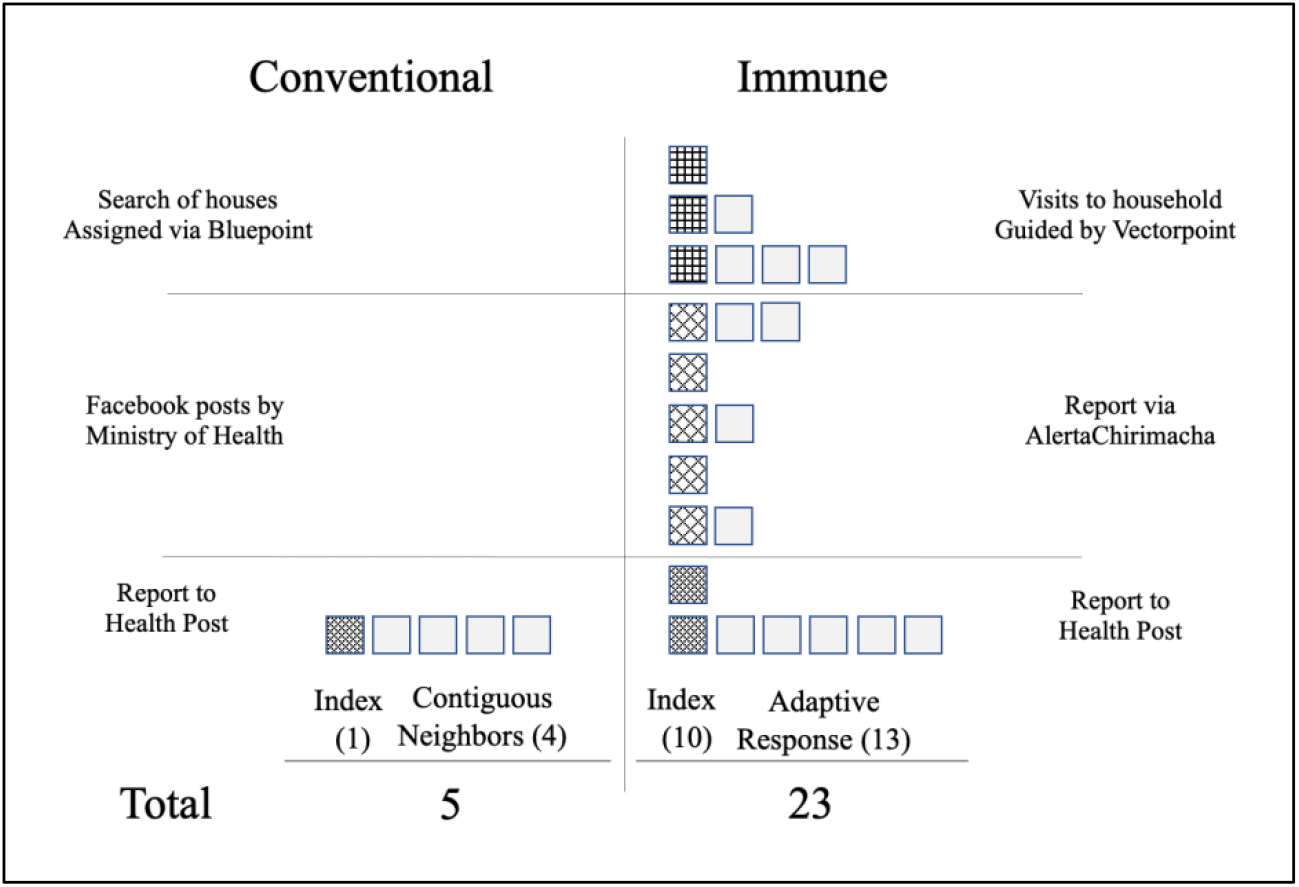
Infestations detected, by modality of detection, under the immune and conventional surveillance arms. *Immune modalities*: AlertaChirimacha includes reports via Whatsapp, Facebook & text. Autonomous search includes inspections of households and visits conducted with the Vectorpoint application, and reports to health posts with or without an insect. *Conventional Modalities* include Facebook posts to the whole city, Search of households assigned through the control app. Reports to the Health posts are only allowed with an insect.

The amount of effort differed between the two arms, mainly due to the activation of the immune arm following detection of infested households. In the immune arm, 837.7 person days were expended by technicians visiting households guided by the Vectorpoint app. An additional 199.5 person days were expended in the adaptive response initiated around the 10 index infestations, and insecticide application to the 23 infested households and immediate neighbors took 48 person days, for a total of 1085.2 person days.

In the control arm, technicians spent 854.2 person days inspecting households assigned to them via the control app. No infested households were detected through the active search. Two additional days were necessary to inspect the neighbors of the index house, and 8 person days were expended in indoor residual insecticide application to infested households and their immediate neighbors for a total of 864.2 total person days of effort.

## Discussion

We challenged the conventional vector control paradigm, formally, against an alternative approach inspired by the adaptive immune system, and the new intervention quickly outperformed the old. Even when fortified with a dozen years of prior data, computational power, and spatial models rarely available to vector control agencies, and the same experienced staff as the intervention arm, the conventional protocols only succeeded in uncovering a single focus of vector infestation of 5 infested houses. Over the same period, the immune arm uncovered 23 infested households in 10 foci spread across 9 sectors of the city.

Urban vector control requires surveillance and control at various scales, as insects spread among houses, blocks, neighborhoods, and cities. The dream of conventional control has long been scalability– the ability to establish a set of protocols that, in the words of Anna Tsing, ‘can expand–and expand, and expand–without rethinking basic elements’. (28) Such sentiments are common in the expansion of demonstration projects into large programs, (29) and, in the early planning of the Southern Cone Initiative to eliminate *Triatoma infestans*, in the focus on normalization of strategies across member states. (30)

The immune system always works across scales. For the most part, the same system that protects a mouse also protects a moose–but it is not scalable, it cannot be effortlessly ported from one place to another, nor expanded over ever-larger geographies. Our protocols in the immune arm allowed technicians to make decisions based on information that would never fit into a statistical model. Such freedom is critical: as the insects are driven towards elimination the predictable infestations are detected and eliminated, leaving behind the idiosyncratic. The immune arm also relied heavily on personal communication and relationships–whether through AlertaChirimacha or door-to-door communication–which cannot be easily scaled. The need for such tailoring of vector surveillance and control is increasingly acknowledged in Chagas disease control, (31) and more broadly, calls for normalization have been replaced by calls for greater attention to local ecologies, geographies, and politics. (32)

Descriptions of the immune system itself are rife with analogy and metaphor. T-cells and pathogens were once described ‘at war’ with one another, (33) and later divided into ‘self’ and ‘nonself’ as a nation with a border, one side of which is in need of protection from the other. (34) The self/nonself analogy has fallen out of favor with increased recognition of the importance of microbiota–a slew of organisms which are not easily, nor consistently, assigned as self or foreign. (35) Understanding of the immune system, and metaphors used to convey that understanding, will continue to evolve, and re-conceptualizations may lead to new ideas about how to control vectors and other disease agents.

For Chagas disease vector surveillance, community reporting is often more sensitive than active search by trained personnel. (36,37) In the immune arm, vector control personnel did not insist on entering homes, but rather focused on communication and dissemination of materials to facilitate reports. The conventional approach relies on quotas—a certain number of homes must be searched. For the immune approach to be formally adopted, the quota system, which sits within a framework of pay for performance, (38–40) would need to be greatly modified or de-implemented. Quotas have a terrible history in Peru—sterilization programs were enacted on a quota system, leading to widespread abuses of women in the 1990s as health personnel conducted sterilizations by force, or without proper consent, as they struggled to reach quotas. (40–43) The quotas for entering and inspecting homes are less nefarious, but still lead to inefficient and irrational behaviors.

Our study had several limitations. Our protocol originally relied heavily on community-based approaches. Community Health Workers were to be the CD4+ T-helper cells, monitoring their neighborhoods and ‘clonally expanding’ by training additional community health workers upon detecting a vector. Perhaps, had COVID-19 and the related lockdowns not prevented the involvement of community health workers, they could have played a pivotal role in the new system.

We shifted our planned communication and clonal expansion from CHWs to social media. In doing so we introduced additional contamination into our study design, as we were not able to target social media posts on a consistent spatial scale around an infestation, and, of course, could not prevent the posts from propagating into areas assigned to the control arm. Indeed, the Ministry of Health reposted our posts regularly. While we were not able to prevent this contamination, we may have lessened it by targeting our posts by including the names of the neighborhoods affected. The contamination would have biased our results toward the null. The formalism of a randomized trial also broke down during the pandemic. The interim analysis we present here was planned for the end of the third year of the trial, and we have conducted it as pre-specified at the end of the third year. But we have less than three years of data on hand.

We have presented here promising results on a new surveillance system, modeled after the mammalian immune system, to control a single insect in a city. But the immune system does not deal with one pathogen at a time, and cities certainly do not deal with infectious agents in isolation. The real power of the analogy may be revealed as it is expanded to address multiple competing threats. To do so may require de-implementation of the old conventional systems to free up resources for a more adaptive and flexible response. It remains unclear whether such a major change in approach will be palatable to control agencies, but the potential is great.

## Data Availability

All data produced in the present work are contained in the manuscript as supplemental information

